# Identifying hidden Zika hotspots in Pernambuco, Brazil: A spatial analysis

**DOI:** 10.1101/2021.03.15.21253657

**Authors:** Laís Picinini Freitas, Rachel Lowe, Andrew E. Koepp, Sandra Valongueiro Alves, Molly Dondero, Letícia J. Marteleto

**Affiliations:** Population Research Center, University of Texas at Austin, Austin, Texas, United States of America; Centre on Climate Change and Planetary Health, London School of Hygiene & Tropical Medicine, London, United Kingdom; Centre for Mathematical Modelling of Infectious Diseases, London School of Hygiene & Tropical Medicine, London, United Kingdom; Department of Human Development and Family Sciences, University of Texas at Austin, Austin, Texas, United States of America; Post-graduation Program of Public Health, Centro de Ciências da Saúde, Universidade Federal de Pernambuco, Recife, Pernambuco, Brazil; Department of Sociology, American University, Washington, D.C., United States of America; Department of Sociology, University of Texas at Austin, Austin, Texas, United States of America

## Abstract

Northeast Brazil has the world’s highest rate of Zika-related microcephaly. Yet, in this hard-hit region, traditional case counts of Zika cannot accurately describe Zika risk. Reporting of Zika cases only became mandatory after its association with microcephaly in neonates, when the Zika epidemic was already declining in the region. To advance the study of the Brazilian Zika epidemic and its impacts, we identified hotspots of Zika in Pernambuco state, Northeast Brazil, using *Aedes*-borne diseases (dengue, chikungunya and Zika) and microcephaly data. We used the Kulldorff’s Poisson purely spatial scan statistic to detect low- and high-risk clusters and combined the results to identify the municipalities most affected by the Zika epidemic. Municipalities were classified as hotspots if they were part of any high-risk cluster, and classified according to a gradient of Zika burden during the epidemic, considering the strength of the evidence. In Pernambuco, officials confirmed 123,934 dengue cases, 167 Zika cases, and 32,983 chikungunya cases between 2014-2017, and 800 microcephaly cases between 2015-2017. We identified 26 *Aedes*-borne diseases clusters (11 high-risk), and 5 microcephaly cases clusters (3 high-risk). Combining the results, sixty-three out of 184 municipalities were identified as hotspots for Zika. The northeast of Pernambuco and the Sertão region were hit hardest by the Zika epidemic. The first is the most populous area, while the second has one of the highest rates of social and economic inequality in Brazil. The identification of Sertão as a Zika hotspot was only possible because the clusters results were combined. The under-reporting of acute infectious diseases is expected to be higher in poor areas. Therefore, using only *Aedes*-borne data does not correctly identify the high-risk areas. We successfully identified hidden Zika hotspots using a simple methodology combining *Aedes-*borne diseases and microcephaly information.

## Introduction

Accurately assessing a community’s disease risk is a major goal in public health research and is critical to the study of epidemics and their consequences. When new viral diseases emerge, this task is challenging, as the reporting of cases may only be established after widespread transmission has occurred. When the Zika virus (ZIKV) reached Brazil in 2014, mandatory reporting of cases did not begin until 2016, at least a full year into the epidemic [1,2].Given the difficulty of accurately assessing Zika burden during the epidemic and the lack of data, identifying areas that were hardest-hit remains a challenge.

ZIKV, a flavivirus transmitted by the *Aedes* mosquitoes, caused its first large outbreak in the island of Yap (Federated States of Micronesia) in 2007 [3]. According to genomics analyses, ZIKV was likely introduced in Brazil in the state of Pernambuco, possibly during 2013, and may have disseminated from there to other regions and even to other countries [4,5]. Zika was considered a benign disease until October 2015, when an unusual increase in the number of neonates with microcephaly was detected in Pernambuco, Northeast Brazil. Microcephaly and other congenital malformations were later associated with ZIKV infection during pregnancy, and the congenital Zika syndrome was first described [1]. Between January 2015 and November 2016, 1,950 infection-related microcephaly cases were confirmed in Brazil, of which 1,487 (76.3%) were in the Northeast region [2]. By the end of the Zika epidemic, nowhere else in the world had microcephaly rates as high as those observed in Northeast Brazil.

The task of identifying high-risk areas for Zika is hampered by some important factors. First, the mandatory reporting of Zika cases to Brazilian health authorities only began in 2016, a full year after the epidemic began, and well after the assumed peak of cases in most of the Northeast region [2,6]. As a result, the number of reported Zika cases in this region and period of time is much lower than the reported cases of congenital microcephaly, despite well-established links between the two [7,8]. In the absence of Zika data for 2014 and 2015, the number of microcephaly cases likely helps to identify areas that experienced Zika outbreaks. It is also important to note that microcephaly cases, given the severity of the condition, are much less likely to be under-reported than Zika cases. In a previous study the incidence of Zika by municipality in Brazil was estimated from the microcephaly cases rate [7]. However, whether the number of microcephaly cases depends only on the Zika incidence is debatable, and other factors – not considered in the aforementioned study – may be acting to modify the risk of this congenital malformation [9–11].

Information on the distribution of other arboviruses endemic in Brazil, namely dengue and chikungunya, can also serve as indicators of Zika incidence during this period. A major factor linking the incidence of Zika, dengue, and chikungunya is the fact that they share the same disease vector, the *Aedes aegypti* mosquito [12]. By virtue of a shared carrier, a greater risk of one arbovirus in a given area implies greater risk of another arbovirus. Furthermore, it can be difficult in a clinical setting to distinguish between symptoms of Zika and symptoms of dengue and chikungunya, and cases of one are sometimes mistakenly reported as cases of another [13]. In fact, in the absence of a channel for reporting Zika cases in 2014 and 2015, the government in Pernambuco encouraged medical professionals to report Zika cases as dengue cases [14]. Thus, the distinction between Zika, dengue, and chikungunya during this period is blurred in official records for clinical as well as administrative reasons.

Therefore, we propose that an elevated risk of Zika during and immediately following the epidemic in Brazil (from 2014-2017) may be detected primarily by an increase in the incidence of microcephaly, as well as by increases in the incidence of dengue, chikungunya, and Zika itself. Considering these factors together represents a way to identify which areas were hit hardest by the Zika epidemic and which areas were less affected. This study sought to identify these areas in the state of Pernambuco by using Kulldorff’s spatial scan statistics for identification of low- and high-risk clusters of *Aedes*-borne diseases (dengue, Zika, and chikungunya) and of microcephaly cases,and classifying the municipalities in a gradient of Zika epidemic burden based on the combination of their cluster status.

## Methods

### Study site

Pernambuco is located in Northeast Brazil, and has 183 municipalities divided in five regions: Agreste, Mata, Metropolitan Region of Recife, São Francisco and Sertão (Fig 1). The state is characterized by coastal and marshy terrain, with varying climate conditions ranging from humid tropical (predominant on the coast) and semiarid (predominant in the interior). The population of Pernambuco was 8,796,448 in 2010, with 89.62 inhabitants per km^2^ [15]. The Metropolitan Region of Recife has the highest population density, with 7,039.64 inhabitants per km^2^ in the capital Recife [16]. Pernambuco is a poor and unequal state in Brazil, and one of the most severely affected states in the Zika epidemic, accounting for 16.8% of Brazil’s reported cases of congenital Zika syndrome through the end of 2017 [17].

**Fig 1.**
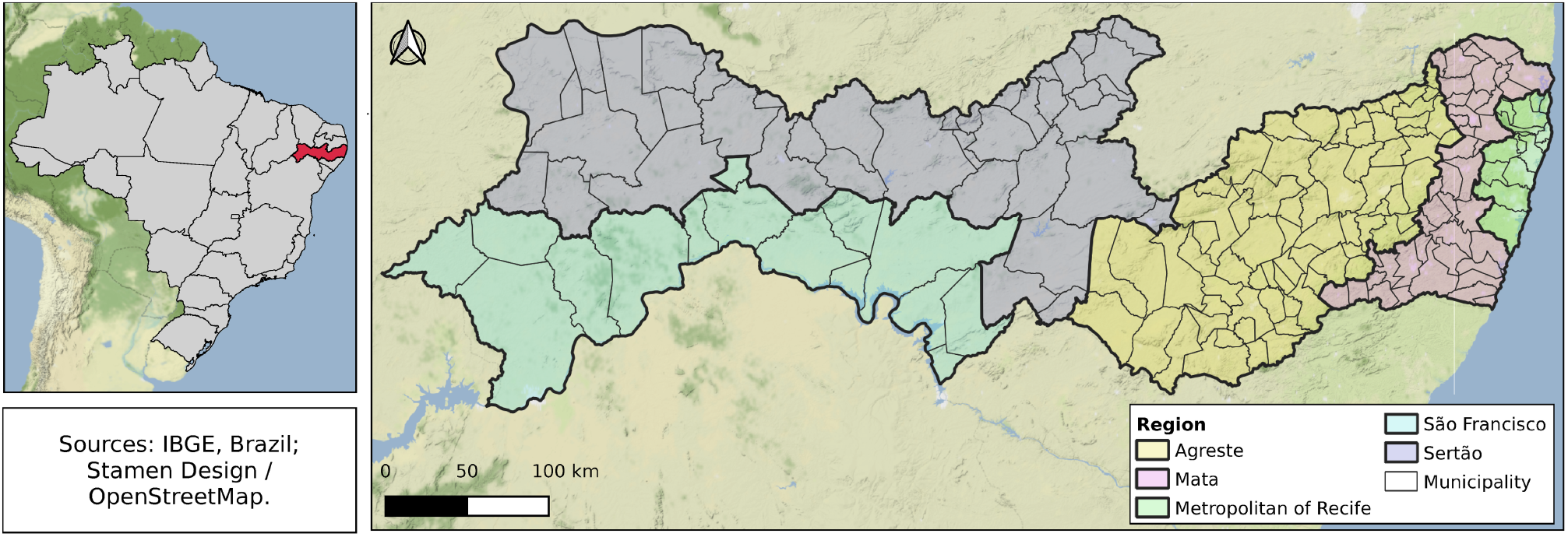
Regions and municipalities of Pernambuco state, Brazil.

### Data

Data on reported confirmed cases of dengue (2013-2017), Zika (2016-2017), and chikungunya (2015-2017) were obtained from the *Sistema de Informação de Agravos de Notificação* [Notifiable Diseases Information System] (SINAN), Brazilian Ministry of Health (ftp://ftp.datasus.gov.br/dissemin/publicos/SINAN/DADOS). The data not publicly available (Zika and chikungunya cases data) were requested from the Ministry of Health at <https://esic.cgu.gov.br/>, by using the Law of Access to Information. We analyzed cases that were confirmed by laboratory or epidemiological criteria, aggregated by municipality of notification and year.

We obtained the anonymized individual records of live births from the Brazilian Ministry of Health’s *Sistema de Informações sobre Nascidos Vivos* [Live Births Information System] (SINASC) from 2013 to 2017. The SINASC data are publicly available at <ftp://ftp.datasus.gov.br/dissemin/publicos/SINASC/>. Microcephaly cases were identified as those with the 10^th^ Revision of the International Statistical Classification of Diseases and Related Health Problems (ICD-10) code “Q02” in any position of the variable corresponding to the classification of congenital anomalies. Then, the data was aggregated by municipality of residence of the mother and year of birth. The complete data were also aggregated to obtain the number of live births by municipality and year.

For the spatial scan statistics, we aggregated the *Aedes*-borne diseases data for the years of 2014-2017, and the microcephaly data for the years of 2015-2017. We chose these years because Zika started causing outbreaks in the state in 2014 and the first Zika-related microcephaly cases were reported in 2015.

Population projections by municipality estimated by Freire *et al*. were used in the analysis [18]. Shapefiles were downloaded at the *Instituto Brasileiro de Geografia e Estatística* [Brazilian Institute of Geography and Statistics] (IBGE) website <https://www.ibge.gov.br/geociencias/organizacao-do-territorio/malhas-territoriais>.

### Statistical analysis

For the exploratory analysis, we calculated the incidence per 100,000 inhabitants for dengue, Zika and chikungunya, and the microcephaly incidence per 10,000 live births, by municipality, for each year and for all years combined. We excluded Fernando de Noronha municipality from the analysis as it is an island roughly 350 km away from the mainland.

We used the packages tidyverse (v. 1.3.0) [19] and ggplot2 (v. 3.3.0) [20] in R (v. 3.6.3) [21] to organize, analyse and visualize the data.

#### Scan statistics

To detect low and high-risk clusters of *Aedes*-borne diseases and microcephaly we used the Kulldorff’s Poisson purely spatial scan statistic [22] for all years combined for i) dengue + Zika + chikungunya (2014-2017), and ii) microcephaly cases (2015-2017).

Purely spatial scan statistics identify clusters through moving circles across space by comparing the observed number of cases to the expected number of cases inside the circle [22]. The municipality was considered as part of the circle if its centroid was located within the circle. The clusters are ordered according to the log-likelihood ratio (LLR), where the cluster with the maximum LLR is the most likely cluster, that is, the cluster least likely to be due to chance [22]. The LLR is calculated as follows:

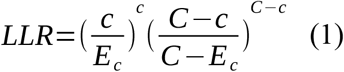

where *c* is the number of cases inside the cluster, *C* is the total number of cases in the state and *E*_*c*_ is the expected number of cases inside the circle. The *E*_*c*_ is calculated by:

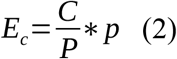

where *P* is the population of the state and *p* is the population inside the cluster.

To assess statistical significance, we performed Monte Carlo simulations (n=999) for each analysis. Clusters were considered to be statistically significant if p-value<0.05. Clusters were restricted to not overlap geographically, to have at least 5 cases, to include a maximum of 50% of Pernambuco’s population at risk, and to have a maximum radius of 50 km.

SaTScan (v. 9.6) software [23] was applied within R (v. 3.6.3) [21], using the package rsatscan (v. 0.3.9200) [24]. The script of the analysis is available at <https://github.com/laispfreitas/PE_satscan>.

#### Zika epidemic burden classification

We combined the results from the scan statistics analysis to identify the municipalities that were most and least affected by the Zika epidemic (Table 1). Municipalities were classified as hotspots if they were part of any high-risk cluster, and as coldspots if they were part of any low-risk cluster and were not part of any high-risk cluster. We also classified the municipalities using a gradient of Zika burden during the epidemic. We considered microcephaly data to provide stronger evidence than *Aedes-*borne diseases because the latter are more likely to be under-reported. Therefore, being part of a high-risk cluster for microcephaly represented a higher Zika burden than being part of a high-risk *Aedes-*borne disease cluster.

**Table 1.** Criteria for classification of municipalities in terms of estimated Zika burden during the epidemic, based on cluster status for microcephaly and dengue, Zika and chikungunya (DZC).

| Zika burden |  | Cluster type |  |
| --- | --- | --- | --- |
| Classification | Gradient | Microcephaly | DZC |
| Hotspots | 5 | High-risk | High-risk |
|  | 4 | High-risk | - |
|  | 3 | High-risk | Low-risk |
|  | 2 | - | High-risk |
|  | 1 | Low-risk | High-risk |
| Neutral | 0 | - | - |
| Coldspots | -1 | - | Low-risk |
|  | -2 | Low-risk | - |
|  | -3 | Low-risk | Low-risk |

## Results

In the state of Pernambuco between 2013 and 2017, there were 128,591 dengue cases, 167 Zika cases, 32,983 chikungunya cases, and 823 microcephaly cases (S1 Table). Of the 167 Zika reported cases, 96 were in Recife. In 2016, 25 municipalities notified at least one case of Zika. Cases of dengue peaked in 2015, while cases of chikungunya peaked in 2016 (Fig 2A). In 2015 the number of microcephaly cases increased dramatically, from 12 in 2014 to 494 cases (Fig 2B).

**Fig 2.**
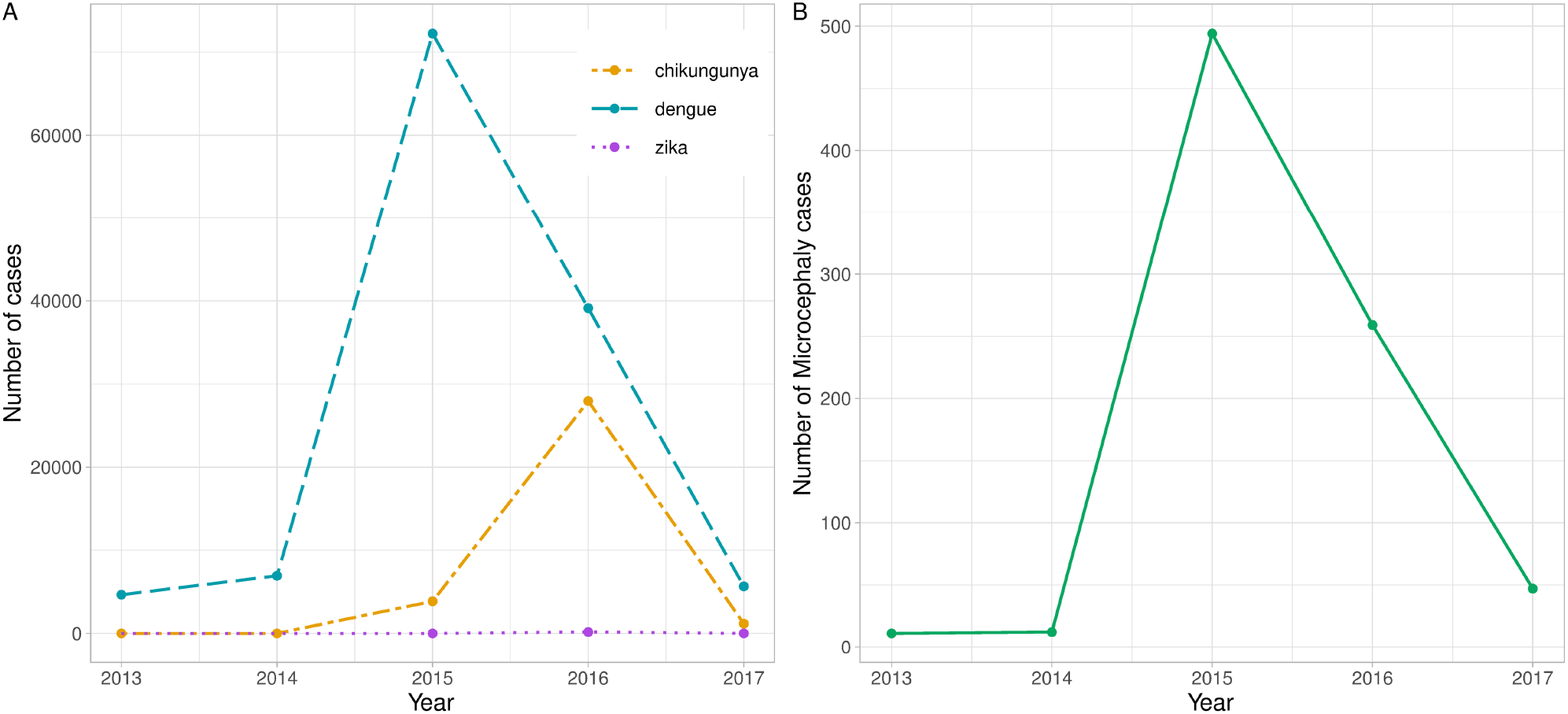
Reported confirmed cases of dengue, Zika and chikungunya (A) and microcephaly in neonates (B) in Pernambuco state, Brazil, by year.

The cumulative incidence of dengue, Zika and chikungunya between 2013-2017 peaked at 12056.6 cases per 100,000 inhabitants, with the highest incidence found in the Agreste region (Fig 3A). In this period, four municipalities had no dengue, Zika, chikungunya or microcephaly cases. Higher microcephaly incidence rates were observed in the Sertão region, peaking at 59.4 cases per 10,000 live-births (Fig 3B). Fifty out of 184 municipalities did not report any cases of microcephaly. The incidence rate per year for each disease and for microcephaly are included in the Supporting Information Material (S1-4 Fig).

**Fig 3.**
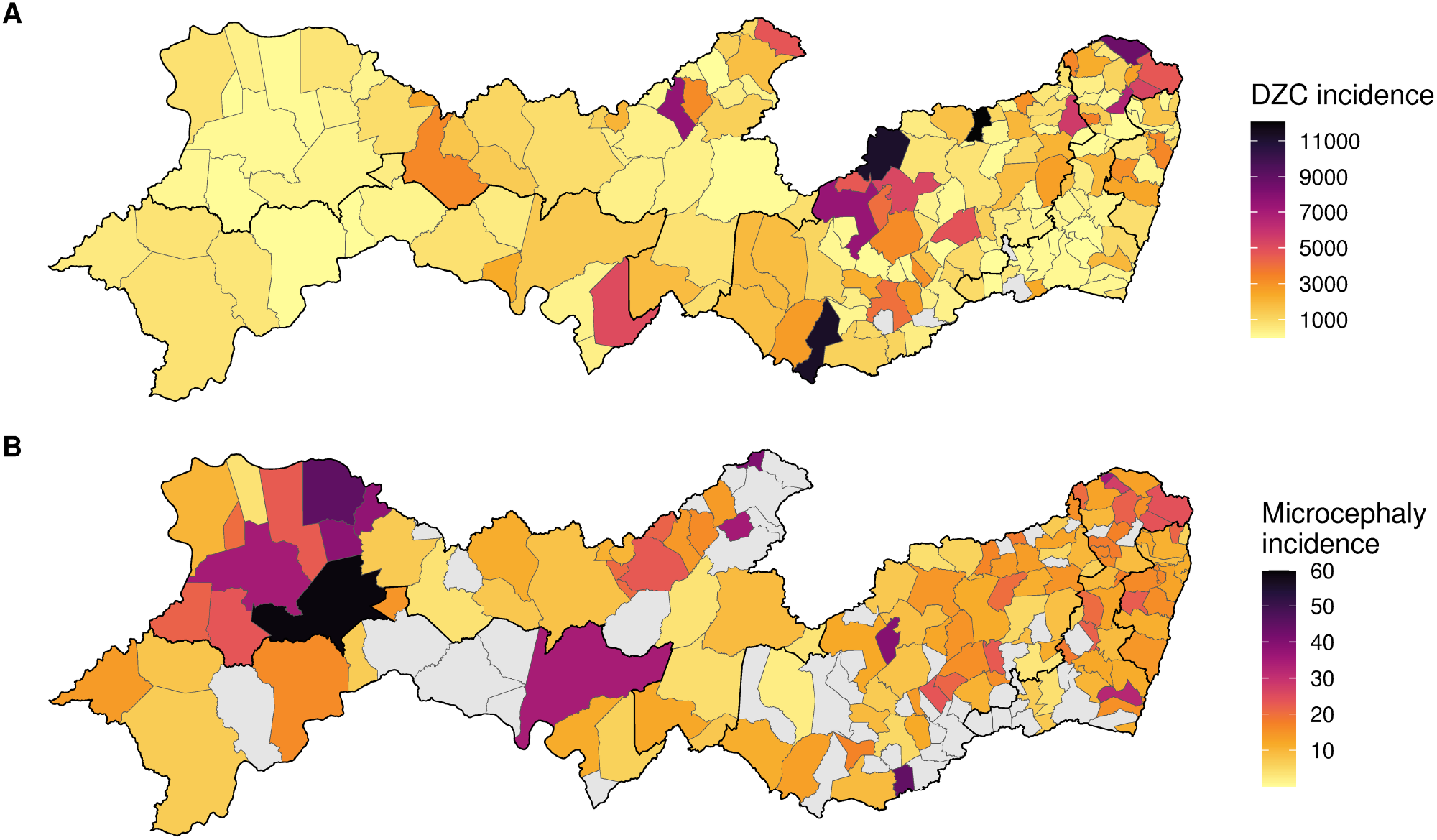
Cumulative incidence of dengue, Zika and chikungunya (DZC) per 100,000 inhabitants (A) and of microcephaly per 10,000 live-births (B), Pernambuco state, Brazil, 2013-2017.

There were 26 clusters of dengue, Zika and chikungunya detected in 2014-2017 using the purely spatial scan statistics, with 11 high-risk and 15 low-risk clusters (S2 Table). The most likely high-risk cluster and the most likely low-risk cluster were both detected in the Metropolitan Region of Recife (Fig 4A). For microcephaly, five clusters were detected in 2015-2017, with three high-risk and two low-risk clusters (S3 Table). The most likely low-risk cluster was detected in the Agreste and Mata regions, and the most likely high-risk cluster in the Northwest of Sertão region (Fig 4B).

**Fig 4.**
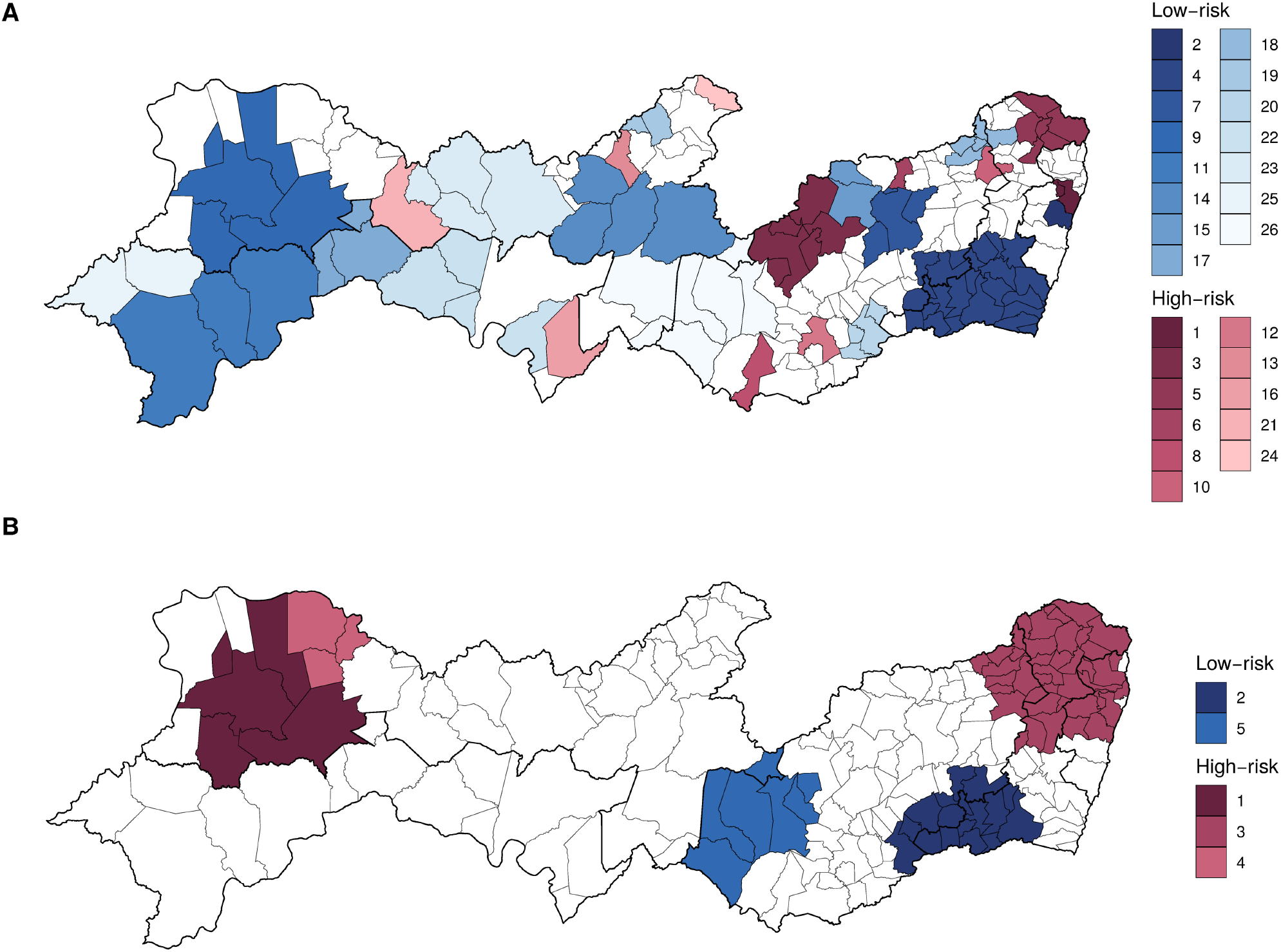
Low and high-risk clusters of dengue, Zika, and chikungunya cases, 2014-2017 (A), and microcephaly, 2015-2017 (B), in Pernambuco state. Clusters are ordered by likelihood ratio.

Combining the results of both scan statistics analyses (*Aedes*-borne diseases and microcephaly), of the 50 municipalities constituting high-risk microcephaly clusters, 10 were also high-risk for dengue, Zika, and chikungunya (Table 2). Of the 24 municipalities constituting low-risk microcephaly clusters, 19 were also low-risk for dengue, Zika, and chikungunya. The names of the municipalities in each category from Table 2 is available in the S4 Table.

**Table 2.** Number and percentage of municipalities by cluster type for dengue, Zika, and chikungunya (DZC) cases (2014-2017), and for microcephaly (2015-2017), Pernambuco state, Brazil.

|  |  | Microcephaly cluster type |  |  | Total |
| --- | --- | --- | --- | --- | --- |
|  |  | High | No cluster | Low |  |
| DZC cluster type | High | 10 (5.4) | 12 (6.5) | 1 (0.5) | 23 (12.5) |
|  | No cluster | 29 (15.8) | 54 (29.3) | 4 (2.1) | 87 (47.3) |
|  | Low | 11 (6.0) | 44 (23.9) | 19 (10.3) | 74 (40.2) |
| Total |  | 50 (27.2) | 110 (59.8) | 24 (13.0) | 184 (100.0) |

In Fig 5 we combined the results from the scan statistics analysis to identify the most and least affected municipalities. The municipalities identified as probable Zika hotspots are depicted in warm colors. Sixty-three out of 184 municipalities were identified as hotspots for Zika. Municipalities in the northeast of Pernambuco state and in the Sertão region were hardest-hit by the Zika epidemic (Fig 5).

**Fig 5.**
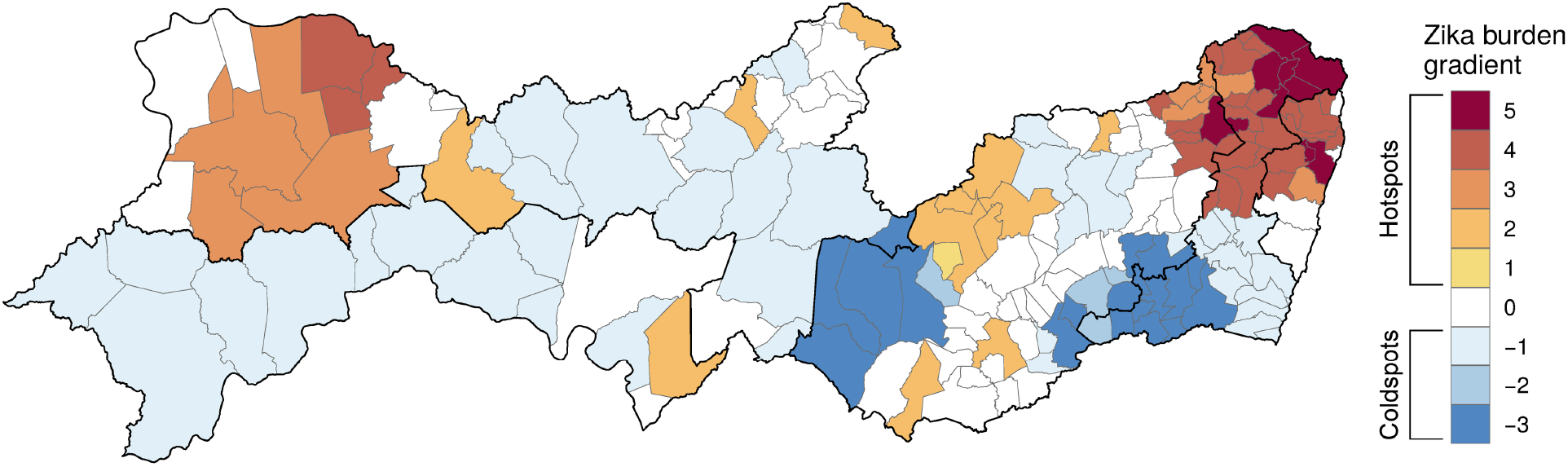
Estimated Zika burden classification by municipality, 2014-2017, Pernambuco state, Brazil.

## Discussion

Mandatory reporting of confirmed Zika cases came late in the epidemic in Brazil, hindering reliable identification of areas of high risk of Zika infection. Further issues with under-reporting also prevented reliable identification of high-risk areas for Zika in the country most affected by the epidemic. To address these issues, we identified spatial clusters of Zika, dengue, and chikungunya – three arboviruses that share the same disease vector, the *Ae. aegypti* mosquito – and of microcephaly in neonates cases to identify hidden Zika hotspots in the state of Pernambuco, one of the most affected by the epidemic.

The two high-risk microcephaly clusters were identified on opposite sides in the state, in the northeast – including parts of the Metropolitan Region of Recife, Mata and Agreste – and in a more western part of the Sertão region. A recent study estimated the spatiotemporal distribution of microcephaly in Pernambuco using a conditional autoregressive model and found high microcephaly prevalences also in the middle portion of the state [28]. In addition to having used a different methodology, the authors used data from a different source, the *Registro de Evento de Saúde Pública* [Public Health Event Registry] (RESP) system, explaining the differences in the results. The RESP system was implemented in November 2015 for the notification of cases of microcephaly or any other congenital anomalies. We opt to use SINASC data because it is more robust, as this system implemented in the country for many decades. In addition, the access to RESP data is more restrict.

Of 23 municipalities constituting high-risk clusters for *Aedes*-borne diseases, ten were also high-risk for microcephaly. One possible explanation is that, in these locations, dengue and/or chikungunya were more prevalent than Zika. However, it has been under discussion whether higher Zika incidence always translates into higher microcephaly incidence [9–11]. Microcephaly rates as high as those observed in Northeast Brazil were unprecedented and not observed anywhere else in the world where Zika has knowingly caused large epidemics. It seems that underlying factors may be acting to modify the risk of developing microcephaly given the infection during pregnancy [11,25–27]. Even today, the reason why some regions of Northeast Brazil presented such high Zika-related microcephaly rates remains an open question.

Fifty municipalities constituted high-risk microcephaly clusters, with only ten of these also constituting high-risk clusters for *Aedes-*borne diseases. Because most cases of microcephaly in the region were caused by Zika [29], the pattern of high risk of microcephaly combined with a low risk of dengue, Zika and chikungunya suggests that there was under-reporting of *Aedes*-borne diseases in these municipalities. The under-reporting of acute infectious diseases is usually higher in poorer areas. As a consequence, using only *Aedes*-borne data would bias the identification of Zika high-risk areas. By combining the analyses using such data with analyses using microcephaly data, we successfully identified hidden Zika hotspots. Of note, the identification of Sertão region as a Zika hotspot was only possible because the scan statistics results for dengue, Zika, and chikungunya were combined with the results for microcephaly.

The northeast of Pernambuco and the Sertão region were hit hardest by the Zika epidemic. The first has the state’s highest population density and urbanization rate. Because other arboviruses are more frequent in urban areas, these areas might see magnified risk of Zika. A recent study in Recife has described an association between precarious living conditions and higher microcephaly prevalence [27]. The urban poor in Brazil often live in households and areas that lack solid infrastructure, such as proper plumbing systems and waste disposal, leading to poor environmental hygiene associated with mosquito breeding. The Sertão region has one of the highest rates of social and economic inequality in Brazil and is also characterized by precarious health care access.

The coldspots need to be interpreted with caution. Further studies are needed to understand whether coldspots were identified as such due to under-reporting or because, in fact, the Zika burden was low in these locations. If the latter is true, the population of municipalities classified as Zika coldspots may be at risk for future outbreaks. Seroprevalence studies can contribute to this issue.

Despite its important contributions, this study has some limitations. Due to the awareness surrounding the microcephaly epidemic, it is possible that microcephaly reporting was over-reported abd a proportion of cases was misdiagnosed. To counterbalance this, we used information from the Live Births System, SINASC, instead of the RESP system, as it is more robust and less prone to bias caused by disease awareness. As already mentioned, there is under-reporting and misclassification of *Aedes*-borne diseases cases. To address the latter, we combined the three diseases – dengue, Zika and chikungunya – in the analysis. Different levels of under-reporting across the municipalities are expected, both for *Aedes*-borne diseases and microcephaly cases, and could bias our results. Finally, we did not consider covariates which might help predict spatial hotspots.

This analysis provides a much-needed classification of Zika risk in the state most affected by the epidemic. In doing so, this study provides a foundation for addressing the potential double jeopardy of two successive novel infectious disease outbreaks. Brazil is the country most affected by the Zika epidemic and has been an epicenter of the COVID-19 pandemic, with nearly 10.5 million confirmed cases and more than 250 thousand deaths by February 28, 2021 [30]. The Brazilian population is therefore experiencing two successive outbreaks with reproductive health consequences. There is now evidence that pregnant women have higher chances of developing the severe form of COVID-19 [31]. There is also evidence of increase in stillbirth and preterm delivery during the pandemic [32]. Women at childbearing ages are further affected by the uncertainty and stress associated with a novel infectious disease, which can also be consequential to pregnant women and fetuses [33]. Such double jeopardy will directly impact how Brazilian women experience reproductive health and childbearing for cohorts to come.

Our study provides a foundation for research investigating social and environmental factors associated with Zika risk, advancing understanding of what makes a location a “risky” place. Merging data on socioeconomic characteristics, health surveillance infrastructure, and environmental conditions at the municipality level to our classification scheme represents an important next step in addressing this question. Our study also provides an important basis for analyses that identify risk of illnesses that are historically under-reported, a common issue particularly in low- and middle-income countries. The applied methodology has the potential to be adapted to instances in which a novel disease emerges and where under-reporting is also expected. As an example, excess deaths, influenza-like illness and hospitalizations due to severe acute respiratory illness, could be used to identify and classify high-risk areas for COVID-19.

The identification of high-risk areas for Zika has important research and policy implications. By combining Zika with other arboviruses and microcephaly, our approach offers a broader and potentially more reliable classification scheme for identifying Zika hotspots – information that can be used to inform public health research and policy. Importantly, our analysis identifies areas that might be particularly vulnerable to under-reporting, as suggested by the clusters that had high microcephaly risk but low *Aedes-*borne diseases risk.

## Data Availability

The data underlying the results presented in the study are available at https://github.com/laispfreitas/PE_satscan/

## Acknowledgements

We would like to thank the current and former members of the Decode Zika project: Ana Paula Portella, Irene Rossetto, Karlos Ramos, Kristine Hopkins, Luiz Gustavo Fernandes Sereno, and Ryan Lloyd.

## Funding

This research was supported by the Eunice Kennedy Shriver National Institute of Child Health and Human Development (https://www.nichd.nih.gov/): grant R01HD091257 awarded to LJM (PI: LJM, co-investigator: MD), and grants P2CH042849 and T32HD007081 awarded to the Population Research Center at the University of Texas at Austin. RL was supported by a Royal Society Dorothy Hodgkin Fellowship (https://royalsociety.org/grants-schemes-awards/grants/dorothy-hodgkin-fellowship/).

## Ethics

This study was conducted under Institutional Review Board approval #2018-01-0055 from the University of Texas at Austin and the Brazilian National Commission for Research Ethics (CONEP – *Comissão Nacional de Ética em Pesquisa*) study approval CAAE: 34032920.1.0000.5149.

## Notes

### Competing Interest Statement

The authors have declared no competing interest.

### Author Declarations

This study was conducted under Institutional Review Board approval #2018-01-0055 from the University of Texas at Austin and the Brazilian National Commission for Research Ethics (CONEP - Comissão Nacional de Ética em Pesquisa) study approval CAAE: 34032920.1.0000.5149.

### Summary of Updates

Correction in the funding information.

## References

1. Lowe R, Barcellos C, Brasil P, Cruz O, Honório N, Kuper H, et al. The Zika Virus Epidemic in Brazil: From Discovery to Future Implications. International Journal of Environmental Research and Public Health. 2018;15: 96. doi:10.3390/ijerph15010096

2. de Oliveira WK, de França GVA, Carmo EH, Duncan BB, de Souza Kuchenbecker R, Schmidt MI. Infection-related microcephaly after the 2015 and 2016 Zika virus outbreaks in Brazil: a surveillance-based analysis. The Lancet. 2017;390: 861–870. doi:10.1016/S0140-6736(17)31368-5

3. Kindhauser MK, Allen T, Frank V, Santhana RS, Dye C. Zika: the origin and spread of a mosquito-borne virus. Bulletin of the World Health Organization. 2016;94: 675–686C. doi:10.2471/BLT.16.171082

4. Costa LC, Veiga RV, Oliveira JF, Rodrigues MS, Andrade RFS, Paixão ES, et al. New Insights on the Zika Virus Arrival in the Americas and Spatiotemporal Reconstruction of the Epidemic Dynamics in Brazil. Viruses. 2020;13: 12. doi:10.3390/v13010012

5. Faria NR, Quick J, Claro IM, Thézé J, de Jesus JG, Giovanetti M, et al. Establishment and cryptic transmission of Zika virus in Brazil and the Americas. Nature. 2017;546: 406–410. doi:10.1038/nature22401

6. Magalhaes T, Braga C, Cordeiro MT, Oliveira AL, Castanha PM, Maciel APR, et al. Zika virus displacement by a chikungunya outbreak in Recife, Brazil. PLOS Neglected Tropical Diseases. 2017;11: e0006055.

7. Brady OJ, Osgood-Zimmerman A, Kassebaum NJ, Ray SE, de Araújo Vem, da Nóbrega AA, et al. The association between Zika virus infection and microcephaly in Brazil 2015–2017: An observational analysis of over 4 million births. Myers JE, editor. PLOS Medicine. 2019;16: e1002755. doi:10.1371/journal.pmed.1002755

8. Johansson MA, Mier-y-Teran-Romero L, Reefhuis J, Gilboa SM, Hills SL. Zika and the risk of microcephaly. NEJM. 2016;375: 1–4. doi:10.1056/NEJMp1605367

9. Rodrigues LC, Paixao ES. Risk of Zika-related microcephaly: stable or variable? The Lancet. 2017;390: 824–826. doi:10.1016/S0140-6736(17)31478-2

10. Costa F, Ko AI. Zika virus and microcephaly: where do we go from here? The Lancet Infectious Diseases. 2018;18: 236–237. doi:10.1016/S1473-3099(17)30697-7

11. Jaenisch T, Rosenberger KD, Brito C, Brady O, Brasil P, Marques ET. Risk of microcephaly after Zika virus infection in Brazil, 2015 to 2016. Bull World Health Organ. 2017;95: 191–198. doi:10.2471/BLT.16.178608

12. Kraemer MUG, Reiner RC, Brady OJ, Messina JP, Gilbert M, Pigott DM, et al. Past and future spread of the arbovirus vectors Aedes aegypti and Aedes albopictus. Nature Microbiology. 2019;4: 854–863. doi:10.1038/s41564-019-0376-y

13. Paixão ES, Teixeira MG, Costa M da CN, Barreto ML, Rodrigues LC. Symptomatic Dengue during Pregnancy and Congenital Neurologic Malformations. Emerging Infectious Diseases. 2018;24: 1748–1750. doi:10.3201/eid2409.170361

14. Brito CAA de, Brito CCM de, Oliveira AC, Rocha M, Atanásio C, Asfora C, et al. Zika in Pernambuco: rewriting the first outbreak. Revista da Sociedade Brasileira de Medicina Tropical. 2016;49: 553–558. doi:10.1590/0037-8682-0245-2016

15. IBGE. Panorama Pernambuco. [cited 3 Aug 2020]. Available: https://cidades.ibge.gov.br/brasil/pe/panorama

16. IBGE. Panorama Recife. [cited 3 Aug 2020]. Available: https://cidades.ibge.gov.br/brasil/pe/recife/panorama

17. Ministério da Saúde B. Monitoramento integrado de alterações no crescimento e desenvolvimento relacionadas à infecção pelo vírus Zika e outras etiologias infecciosas, até a Semana Epidemiológica 52 de 2017. Boletim Epidemiológico. 2018;49. Available: https://www.saude.gov.br/images/pdf/2018/fevereiro/20/2018-003-Final.pdf

18. Freire FHM de A, Gonzaga MR, Gomes MMF. Projeções populacionais por sexo e idade para pequenas áreas no Brasil. Revista Latinoamericana de Población. 2019;14: 124–149. doi:10.31406/relap2020.v14.i1.n26.6

19. Wickham H, Averick M, Bryan J, Chang W, McGowan L, François R, et al. Welcome to the Tidyverse. Journal of Open Source Software. 2019;4: 1686. doi:10.21105/joss.01686

20. Wickham H. ggplot2: Elegant Graphics for Data Analysis. Springer-Verlag. 2016. Available: https://ggplot2.tidyverse.org/

21. The R Foundation for Statistical Computing. R. The R Foundation; 2020. Available: https://www.r-project.org/

22. Kulldorff M. A spatial scan statistic. Communications in Statistics - Theory and Methods. 1997;26: 1481–1496. doi:10.1080/03610929708831995

23. Kulldorff M. SaTScan. Available: https://www.satscan.org/

24. Kleinman K. rsatscan: Tools, Classes, and Methods for Interfacing with SaTScan Stand-Alone Software. 2015. Available: https://CRAN.R-project.org/package=rsatscan

25. Carvalho MS, Freitas LP, Cruz OG, Brasil P, Bastos LS. Association of past dengue fever epidemics with the risk of Zika microcephaly at the population level in Brazil. Scientific Reports. 2020;10. doi:10.1038/s41598-020-58407-7

26. Santa Rita TH, Barra RB, Peixoto GP, Mesquita PG, Barra GB. Association between suspected Zika virus disease during pregnancy and giving birth to a newborn with congenital microcephaly: a matched case–control study. BMC Research Notes. 2017;10. doi:10.1186/s13104-017-2796-1

27. Souza WV de, Albuquerque M de FPM de, Vazquez E, Bezerra LCA, Mendes A da CG, Lyra TM, et al. Microcephaly epidemic related to the Zika virus and living conditions in Recife, Northeast Brazil. BMC Public Health. 2018;18. doi:10.1186/s12889-018-5039-z

28. Alexander NDE, Souza WV, Rodrigues LC, Braga C, Sá A, Albuquerque Bezerra LC, et al. Spatiotemporal Analysis of the Population Risk of Congenital Microcephaly in Pernambuco State, Brazil. International Journal of Environmental Research and Public Health. 2020;17: 700. doi:10.3390/ijerph17030700

29. de Araújo Tvb, Ximenes RA de A, Miranda-Filho D de B, Souza WV, Montarroyos UR, de Melo Apl, et al. Association between microcephaly, Zika virus infection, and other risk factors in Brazil: final report of a case-control study. The Lancet Infectious Diseases. 2018;18: 328– 336. doi:10.1016/S1473-3099(17)30727-2

30. WHO. Weekly epidemiological update - 2 March 2021. 2021 [cited 16 Dec 2020]. Available: https://www.who.int/publications/m/item/weekly-epidemiological-update 2-march-2021

31. CDC C for DC and P. CDC updates, expands list of people at risk of severe COVID-19 illness. In: Centers for Disease Control and Prevention [Internet]. 25 Jun 2020 [cited 8 Mar 2021]. Available: https://www.cdc.gov/media/releases/2020/p0625-update-expands-covid-19.html

32. Khalil A, von Dadelszen P, Draycott T, Ugwumadu A, O’Brien P, Magee L. Change in the Incidence of Stillbirth and Preterm Delivery During the COVID-19 Pandemic. JAMA. 2020 [cited 20 Jul 2020]. doi:10.1001/jama.2020.12746

33. Torche F. The Effect of Maternal Stress on Birth Outcomes: Exploiting a Natural Experiment. Demography. 2011;48: 1473–1491. doi:10.1007/s13524-011-0054-z

