## Supplementary material for "Identifying hidden Zika hotspots in Pernambuco, Brazil: A spatial analysis": S1 Table

Supporting Information Material

Laís P Freitas, Rachel Lowe, Andrew E Koepp, Sandra V Alves, Molly Dondero, Letícia J Marteleto

**S1 Table. Cases of dengue, Zika, chikungunya and microcephaly in Pernambuco state, Brazil, by year.**

| Year | Dengue | Zika | Chikungunya | Microcephaly |
|---|---|---|---|---|
| 2013 | 4657 | 0 | 0 | 11 |
| 2014 | 6934 | 0 | 0 | 12 |
| 2015 | 72220 | 0 | 3865 | 494 |
| 2016 | 39116 | 163 | 27947 | 259 |
| 2017 | 5664 | 4 | 1171 | 47 |

**S1 Fig. Dengue incidence (cases by 100,000 inhabitants) by year and municipality in Pernambuco state, Brazil.**

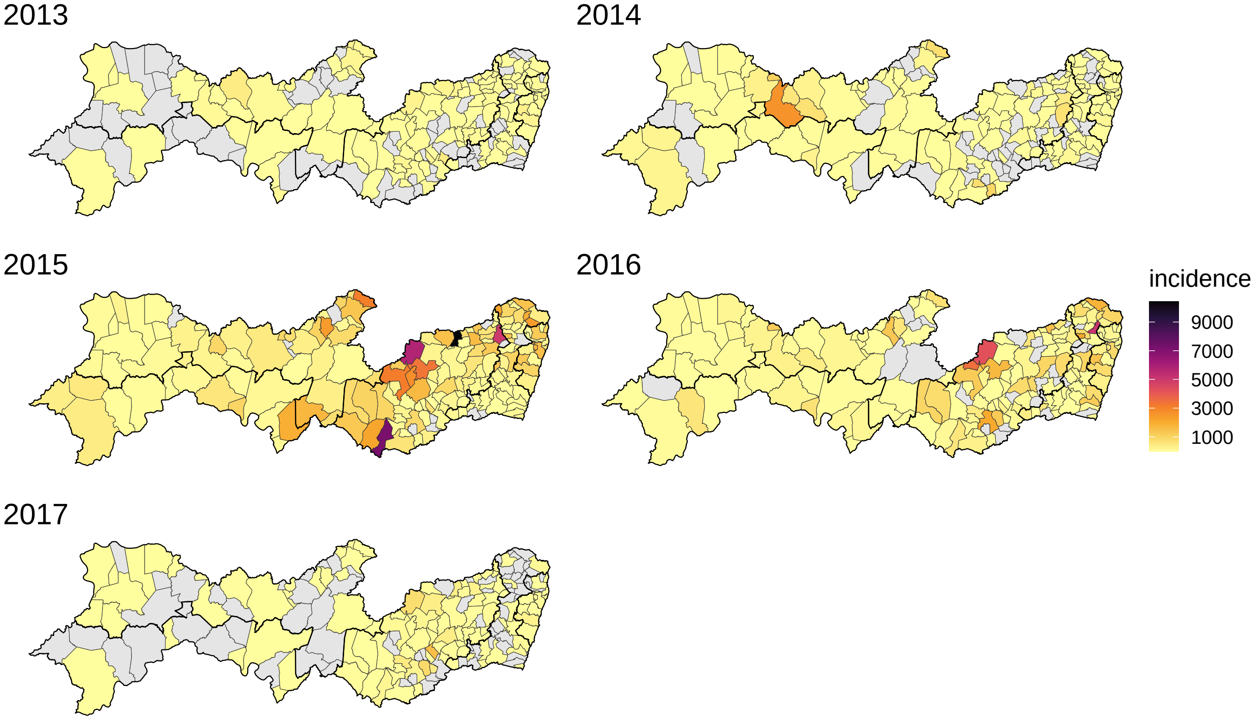

**S2 Fig. Zika incidence (cases by 100,000 inhabitants) by year and municipality in Pernambuco state, Brazil.**

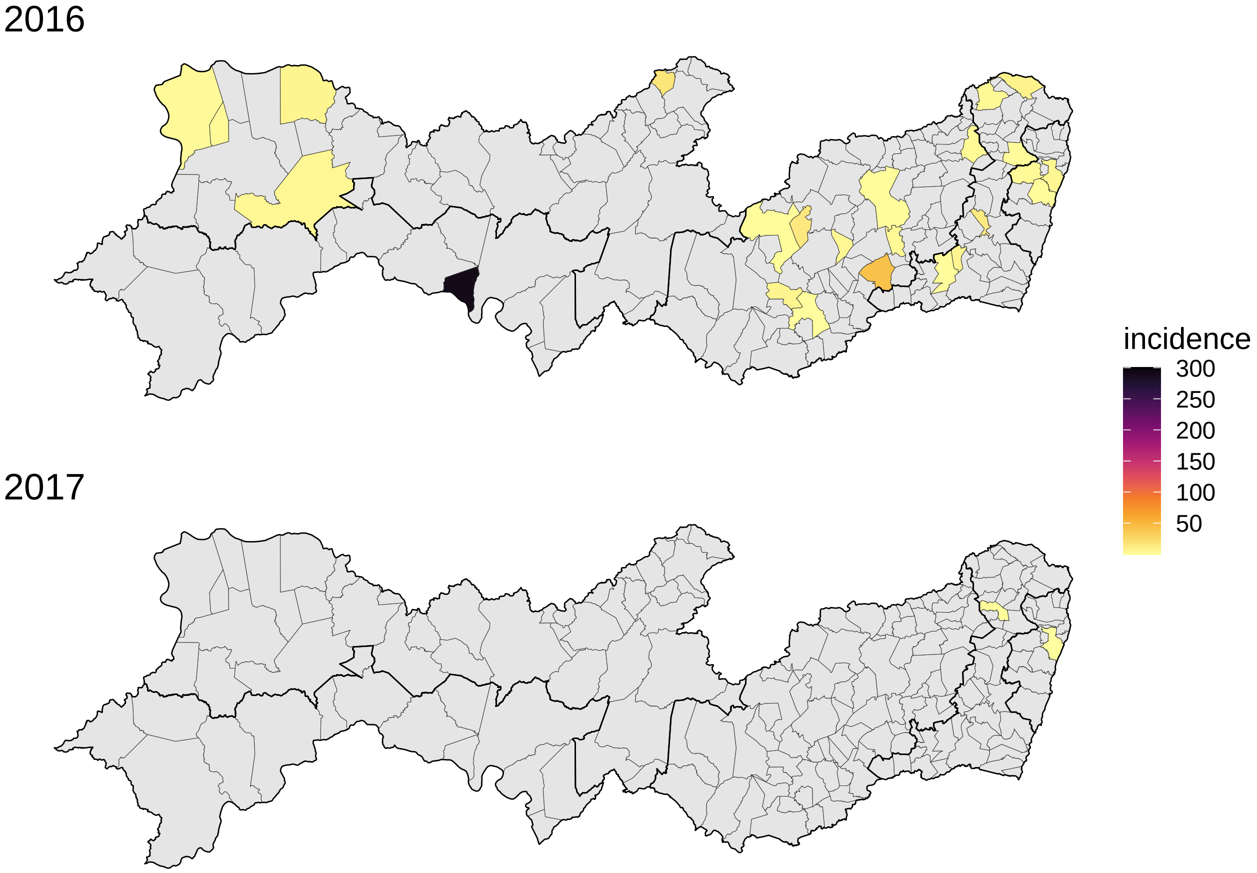

**S3 Fig. Chikungunya incidence (cases by 100,000 inhabitants) by year and municipality in Pernambuco state, Brazil.**

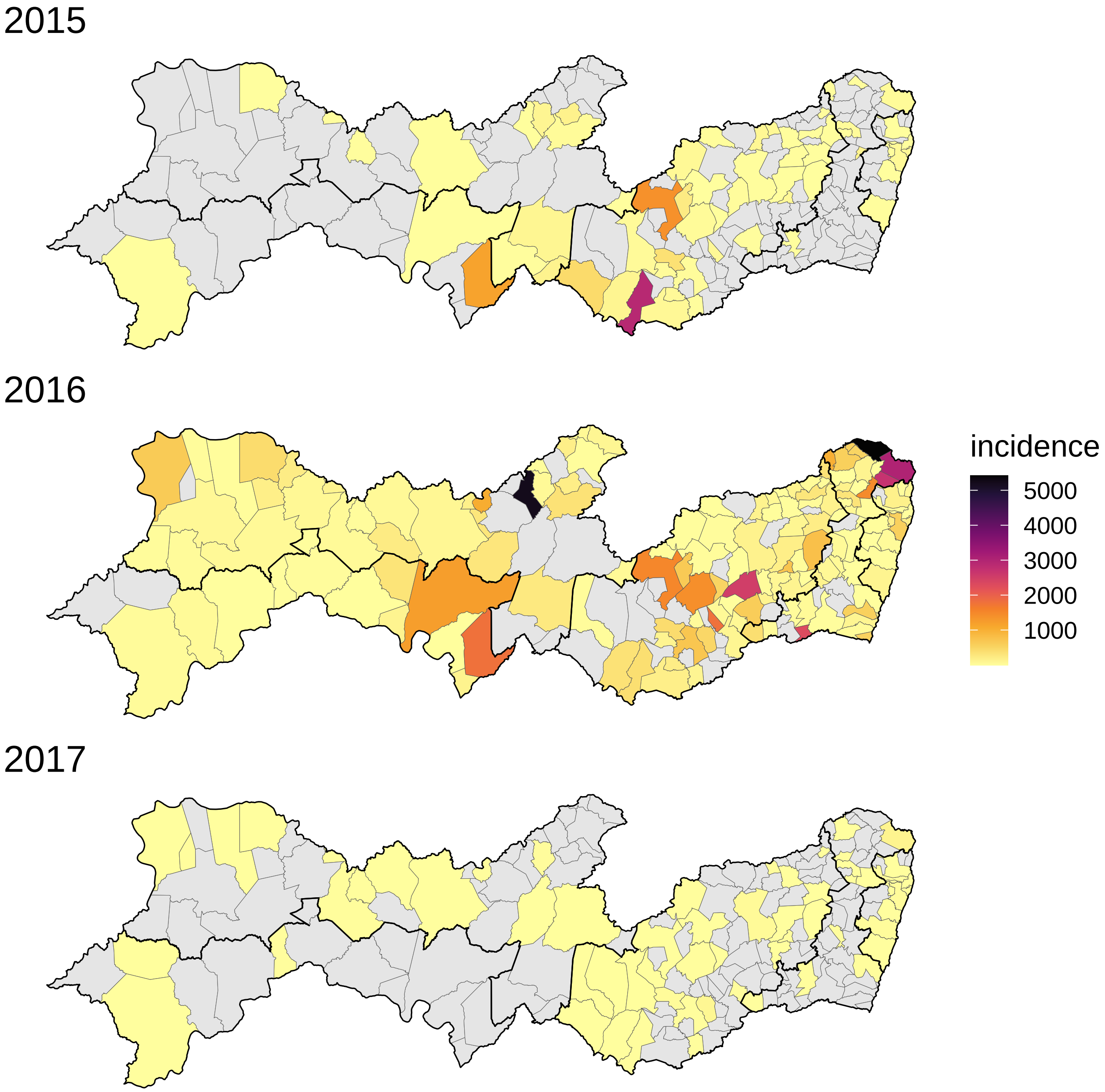

**S4 Fig. Microcephaly incidence (cases by 10,000 live-births) by year and municipality in Pernambuco state, Brazil.**

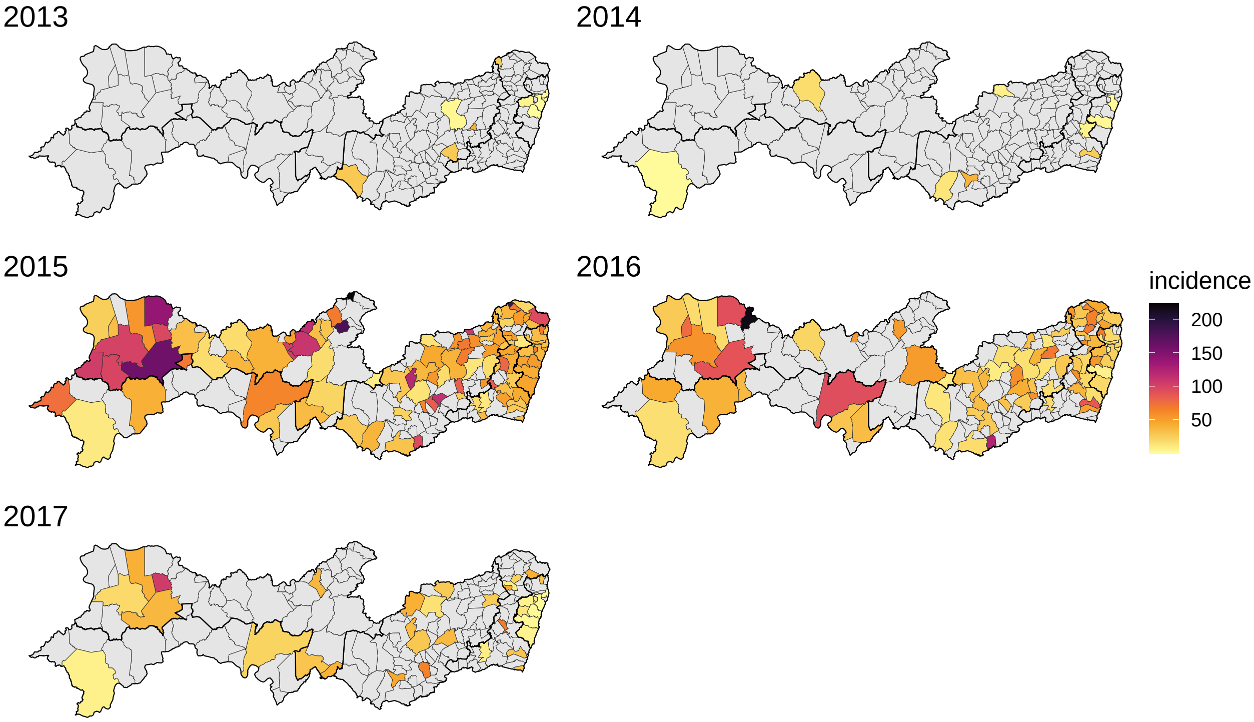

**S2 Table. Dengue, Zika and chikungunya low and high-risk clusters in Pernambuco state, 2014-2017.**

| Cluster | Population | Observed cases | Expected cases | Relative risk | Type |
|---|---|---|---|---|---|
| 1 | 1747169 | 55922 | 29376.1064 | 2.4031938 | high |
| 2 | 677128 | 1358 | 11384.9132 | 0.1116004 | low |
| 3 | 208879 | 12231 | 3511.9889 | 3.6922697 | high |
| 4 | 657905 | 2357 | 11061.7119 | 0.2010899 | low |
| 5 | 207578 | 9712 | 3490.1222 | 2.9001935 | high |
| 6 | 20235 | 2376 | 340.2263 | 7.0754827 | high |
| 7 | 459022 | 2567 | 7717.7882 | 0.3215208 | low |
| 8 | 18877 | 2138 | 317.3967 | 6.8151972 | high |
| 9 | 171289 | 223 | 2879.9707 | 0.0761198 | low |
| 10 | 72429 | 3703 | 1217.7877 | 3.0900289 | high |
| 11 | 414454 | 2824 | 6968.4408 | 0.3943678 | low |
| 12 | 136859 | 5455 | 2301.0857 | 2.4199295 | high |
| 13 | 19066 | 1451 | 320.5705 | 4.5591817 | high |
| 14 | 106585 | 291 | 1792.0734 | 0.1608272 | low |
| 15 | 160206 | 820 | 2693.6388 | 0.3007709 | low |
| 16 | 25246 | 1281 | 424.4762 | 3.0344275 | high |
| 17 | 58542 | 155 | 984.2962 | 0.1566408 | low |
| 18 | 127164 | 854 | 2138.0848 | 0.3961400 | low |
| 19 | 33682 | 14 | 566.3182 | 0.0246342 | low |
| 20 | 58213 | 208 | 978.7672 | 0.2114681 | low |
| 21 | 59649 | 1922 | 1002.9127 | 1.9277699 | high |
| 22 | 74237 | 459 | 1248.1867 | 0.3658806 | low |
| 23 | 142276 | 1292 | 2392.1566 | 0.5362844 | low |
| 24 | 13651 | 638 | 229.5286 | 2.7868678 | high |
| 25 | 37483 | 247 | 630.2169 | 0.3909709 | low |
| 26 | 254480 | 3319 | 4278.7106 | 0.7708595 | low |

**S3 Table. Microcephaly low and high-risk clusters in Pernambuco state, 2015-2017.**

| Cluster | Live Births | Observed cases | Expected cases | Relative risk | Type |
|---|---|---|---|---|---|
| 1 | 8306 | 44 | 16.144339 | 2.8258345 | high |
| 2 | 19677 | 11 | 38.246106 | 0.2776791 | low |
| 3 | 178311 | 409 | 346.582375 | 1.3684801 | high |
| 4 | 2165 | 16 | 4.208102 | 3.8593769 | high |
| 5 | 10021 | 4 | 19.477777 | 0.2013691 | low |

**S4 Table. Name of the municipalities by cluster type for dengue, Zika, and chikungunya cases (2014-2017), and for microcephaly (2015-2017), Pernambuco state, Brazil.**

| | | Microcephaly | | |
|---|---|---|---|---|
| | | **High** | **Low** | **No** |
| **DZC** | **High** | Aliança, Camaragibe, Condado, Goiana, Itambé, Itaquitinga, Lagoa Do Carro, Limoeiro, Recife, Tracunhaém | Alagoinha | Belo Jardim, Carnaíba, Garanhuns, Iati, Itapetim, Jataúba, Pesqueira, Poção, Salgueiro, Sanharó, Tacaratu, Vertentes |
| | **Low** | Bodocó, Bom Jardim, Jaboatão Dos Guararapes, Machados, Orobó, Ouricuri, Parnamirim, Santa Cruz, São Vicente Ferrer, Trindade, Vicência | Água Preta, Arcoverde, Belém De Maria, Bonito, Buíque, Canhotinho, Catende, Itaíba, Jaqueira, Joaquim Nabuco, Jurema, Lagoa Dos Gatos, Maraial, Palmares, Pedra, São Benedito Do Sul, São Joaquim Do Monte, Tupanatinga, Xexéu | Afrânio, Amaraji, Angelim, Barra De Guabiraba, Barreiros, Belém Do São Francisco, Betânia, Brejo Da Madre De Deus, Cabrobó, Carnaubeira Da Penha, Caruaru, Cortês, Custódia, Dormentes, Escada, Flores, Gameleira, Ibimirim, Itacuruba, Lagoa Grande, Manari, Mirandiba, Orocó, Palmeirina, Petrolândia, Petrolina, Primavera, Riacho Das Almas, Ribeirão, Rio Formoso, Santa Cruz Do Capibaribe, Santa Maria Da Boa Vista, São Caitano, São José Da Coroa Grande, São José Do Belmonte, Serra Talhada, Sertânia, Sirinhaém, Solidão, Tabira, Tamandaré, Terra Nova, Toritama, Verdejante |
| | **No** | Abreu E Lima, Araçoiaba, Buenos Aires, Camutanga, Carpina, Casinhas, Chã De Alegria, Chã Grande, Exu, Feira Nova, Ferreiros, Glória Do Goitá, Granito, Igarassu, Itapissuma, João Alfredo, Lagoa De Itaenga, Macaparana, Moreno, Nazaré Da Mata, Passira, Paudalho, Paulista, Pombos, Salgadinho, São Lourenço Da Mata, Moreilândia, Timbaúba, Vitória De Santo Antão | Cupira, Panelas, Quipapá, Venturosa | Afogados Da Ingazeira, Agrestina, Águas Belas, Altinho, Araripina, Bezerros, Bom Conselho, Brejão, Brejinho, Cabo De Santo Agostinho, Cachoeirinha, Caetés, Calçado, Calumbi, Camocim De São Félix, Capoeiras, Cedro, Correntes, Cumaru, Floresta, Frei Miguelinho, Gravatá, Ibirajuba, Iguaracy, Inajá, Ingazeira, Ipojuca, Ipubi, Ilha De Itamaracá, Jatobá, Jucati, Jupi, Lagoa Do Ouro, Lajedo, Olinda, Paranatama, Quixaba, Sairé, Saloá, Santa Cruz Da Baixa Verde, Santa Filomena, Santa Maria Do Cambucá, Santa Terezinha, São Bento Do Una, São João, São José Do Egito, Serrita, Surubim, Tacaimbó, Taquaritinga Do Norte, Terezinha, Triunfo, Tuparetama, Vertente Do Lério |
